# Returning to nursing during the COVID-19 pandemic: experiences and needs of re-entering nurses

**DOI:** 10.1101/2021.02.11.21251571

**Authors:** Sofie A. Noorland, Trynke Hoekstra, Maarten O. Kok

## Abstract

**Aim:** Assessing the needs and experiences of re-entering nurses during the COVID-19 pandemic.

**Background:** During the COVID-19 outbreak in the Netherlands, thousands of former nurses have returned to nursing to support healthcare staff. After a period of absence and with little time to prepare, these former nurses re-entered during a challenging, uncertain and rapidly evolving pandemic. Little is known about the experiences and needs of these re-entering nurses.

**Design:** Qualitative study

**Methods:** We conducted semi-structured interviews with 20 purposively selected nurses who had re-entered nursing during the first wave of the COVID-19 pandemic in the Netherlands. Interviews were transcribed verbatim and analysed via thematic content analysis. This study followed the COREQ guidelines.

**Results:** Participants mentioned that a lack of a clear job description led to unclarity about the kind of tasks that re-entering nurses were expected and allowed to perform. This unclarity was especially notable in the newly established COVID-19 departments. Re-entering nurses mentioned to wish for an easily accessible mentorship structure and an individualised and practical training program. Re-entering nurses felt supported by a positive team dynamic, which was shaped by the sense of urgency and relevance of their work and helped them deal with stressful experiences.

**Conclusion:** The results indicate that a rapid and safe return to nursing during a pandemic could be facilitated by: a clear description of roles and responsibilities; an individualised assessment determining the competences and knowledge disparities of re-entering nurses; practical training focussing on competencies needed during a pandemic; and a collaborative mentorship structure to guide re-entering nurses.

**Relevance to clinical practice:** The rapid recruitment of former nurses to mitigate an acute shortage of qualified nurses could play a vital role during a pandemic. To deploy these nurses effectively, safely and sustainably, it is important to address the needs of these re-entering nurses.

**What does this paper contribute towards the wider global clinical community?:**

- This research showed the need to prepare a flexible individualised training programme which could support re-entering nurses during crisis situations, such as a pandemic.
- A responsive mentorship structure helps to provide support to re-entering nurses in a dynamic, uncertain and rapidly evolving situation.
- In a rapidly evolving situation, it is essential to continue to create clarity about the roles and responsibilities of re-entering nurses.

## Introduction

The worldwide shortage of nurses, which is estimated to be over six million, is further pressurized by the COVID-19 pandemic (World Health Organisation (WHO), 2020). Many patients with COVID-19 require hospitalisation and/or specialised care, which is heavily straining healthcare capacity and leads to a rapid increase in the need for qualified nurses (Huang et al., 2020; Zhu & Zhang et al., 2019).

In the Netherlands, the need to rapidly deploy more nurses became even before COVID-19 was declared a pandemic in March 2020. Before the start of the pandemic, the Dutch Ministry of Health (MOH) was anticipating that the shortage of health workers would rise to over 80,000 by 2022 (ministerie van Volksgezondheid, Welzijn en Sport (VWS), 2019). To rapidly expand the number of nurses who could be deployed during the pandemic, the Dutch MOH supported the recruitment of former nurses willing to re-enter into nursing (VWS, 2020a; Bruins, 2020). Approximately, over five thousand former nurses re-entered in different settings of the Dutch healthcare system at the height of the first wave (Wapenaar, 2020). Three important challenges of this process requiring further study were identified: nursing during a new, emerging pandemic, little preparation time and re-entering after a (long) period of absence.

Nursing during a rapidly evolving pandemic is challenging. Health workers in the frontlines are subjected to multiple hazards, such as prolonged working hours, shortages of personal protective equipment (PPE), and a higher risk of infection, due to the increased exposure to infected patients (Lai et al., 2020; Zhu, Xu et al., 2020). Moreover, early research during the beginning of the pandemic in Wuhan China reported nurses experiencing anxiety, depression and stress (Zhu, Xu et al., 2020). Comparable findings have been reported during previous MERS, Ebola and SARS outbreaks (Khalid et al., 2016; Maunder et al., 2003; Cénat et al, 2020). In contrast, studies have also reported that providing care during a crisis situation, such as the civil war in Guinea-Bissau, can lead to an increase in dedication, focus and morale among staff, which is thought to have contributed to a decrease in infant mortality rates under five in hospitals (Biai et al., 2007).

Additionally, research shows that former nurses returning to health care have indicated to struggle with anxiety and insecurities about their capabilities, changed roles, new technologies, the amount of paperwork, accountability measures and responsibility levels and a negative attitude from other health-care personnel when re-entering after a (long) period of absence (Durand & Randhawa, 2002; Long & West, 2007).

Little is known about how former nurses perceive the process of re-entering and what their needs are in times of a newly emerging and ongoing pandemic. The current pandemic will presumably continue until effective vaccines are widely available and rolled out effectively and new mutations and other pandemics are likely to emerge (Cui et al., 2019; Anderson et al., 2020). Hence, the rapid recruitment of former nurses to mitigate an acute shortage of qualified nurses is likely to be vital in new and evolving pandemics and other health crises. To learn how former nurses re-entering during the COVID-19 pandemic can be best supported, this qualitative study aims to assess their experiences and needs during the first wave of the COVID-19 pandemic in the Netherlands.

## Methods

We conducted a qualitative study, using an interpretative phenomenological approach, which emphases the lived experiences of participants in the context of their ‘real world’ (Green & Thorogood, 2018). To present the methods section, we used the Consolidated criteria for Reporting Qualitative research (COREQ) (c.f. Supplementary File 1)

### Study population

Purposive sampling of re-entering nurses employed in different settings in healthcare enabled the comparison in experiences and needs within various settings. Via social media platforms, re-entering nurses were approached and asked to participate. Additionally, four participants were recruited through snowball sampling.

Re-entering nurses were included if they had not worked on a nursing ward for at least one year and re-entered between March 2020 and June 2020 to make sure that the re-entering process indeed took place in the context of the first wave of the COVID-19 pandemic. Participants had to be employed within the following settings: hospitals, rehabilitation centres, home care, nursing home settings or newly established COVID-19 departments within nursing home settings. A total of 20 participants were included for this research.

### Data collection

Data collection was done through one-on-one in-depth semi-structured interviewing. A semi-structured interview guide was based upon concepts identified in the literature (appendix 1) as well as intuition and consisted of themes with leading questions and examples for prompts (appendix 2). Interviews lasted 56-110 (Med 85,5) minutes and were conducted through Skype because of social distancing regulations. One interview was conducted by phone because of technical issues. Interviews were audio-recorded and transcribed afterwards.

Memos were made during the interview process and were afterwards directly supplemented with short summaries of the interviews.

### Data analysis

Data collection and analysis were an iterative process. The analysis started after the first interview, hence resulted in a cyclical process in which the researcher moves back and forth between data and analysis (Green & Thorogood, 2018). The steps and decisions that were made during the study were described in an audit trail. A thematic content analysis was used to analyse data. The first step in the process was familiarisation with the data, done through re-listening to tapes and reading through transcripts and interview summaries (Green & Thorogood, 2018). Secondly, data of five interviews were coded, and regularities were inductively categorised in themes. The emerging coding scheme was then applied to all data (appendix 3). Finally, data from all cases are organised horizontally across the themes. MAXQDA2020 (VERBI Software, 2019) was used for organisation and coding. Data saturation on the main themes was reached after 13 interviews.

### Research ethics

The study proposal was checked and approved by the ethical committee of the Faculty of Science of the Vrije Universiteit Amsterdam (https://beta.vu.nl/nl/onderzoek/research-ethics-review/index.aspx). According to the committee, the study was not subject to the Medical Research Involving Human Subjects Act (WMO). Informed consent was obtained from all participants prior to the interview.

The researcher conducting the interviews (SAN) has a background in nursing and was employed as a nurse at a COVID-19 department during the conduct of this study. A diary was kept as a reflexivity tool to avoid personal assumptions and preconceptions. Participants were aware of the background of the researcher and the study aims.

## Results

### Characteristics

Characteristics of the 20 participants are described in table 1. Most participants re-entered care within a period of days to two weeks after applying. At the time of the interview, most nurses had re-entered in nursing for at least a month. Most of the former nurses who were able to retain their BIG-registration (appendix 4) still worked in a profession closely related to individual healthcare (e.g. nursing management or nursing teaching positions). Two returners were able to retain a BIG-registration due to the adjustments in the Dutch regulations that temporarily revoked the expiry of the BIG-registration.

**Table 1.** Characteristics of respondents (n=20)

|  | n |
| --- | --- |
| Male | 4 |
| Female | 16 |
| **Years of experience in nursing (pre-exit)** |  |
| >0 ≤5 years | 2 |
| >5 ≤10 years | 6 |
| >10 ≤15 years | 3 |
| >15 years | 9 |
| **Years since last job in nursing** |  |
| ≥1 ≤5 | 4 |
| >5 ≤10 | 8 |
| >10 ≤15 | 3 |
| >15 | 5 |
| **Weeks back in nursing at moment of interview** |  |
| ≤2 | 1 |
| >2 ≤4 | 2 |
| >4 ≤6 | 5 |
| >6 | 12 |
| **BIG-registered at moment of return** |  |
| Yes | 9 |
| No | 11 |

**Table 2.** Healthcare settings and job positions

|  |  | Setting |  |  |  |
| --- | --- | --- | --- | --- | --- |
|  |  | Nursing home/<br>rehabilitation<br>centre | COVID-19<br>department<br>nursing home<br>setting | Intensive care<br>hospital | General/<br>COVID-19<br>department<br>hospital |
| Position | Independent<br>baccalaureate-<br>educated/Vocationally<br>trained independent<br>registered nurses | 1 | 3 | 3 | 2 |
|  | Nurse assistant<br>staffing | 1 | 1 | - | 1 |
|  | Nurse aide staffing | 2 | - | - | - |
|  | Buddy position | - | - | 5 | 1 |

Participants were placed in different settings and operated in various positions within the Dutch healthcare system. Placements depended on the preferences of participants and the needs of healthcare organisations in the different regions of the Netherlands. All nurses who had experience in working at an intensive care unit (ICU) were positioned at ICUs in hospitals, regardless of the number of years they were absent from nursing. For instance, one former ICU nurse had quit nursing in 1999, yet was still employed to assist at an ICU. All re-entering nurses working on ICUs were confronted with COVID-19 patients. Within nursing home settings, new departments were established to accommodate the increasing number of COVID-19 patients. In two occasions hotels were temporarily converted into COVID-19 units. Three respondents worked in newly established departments for low-complex COVID-19 patients or non-COVID patients with low-complex care demands within hospital settings. These departments were constructed to reduce the pressure on healthcare within hospitals.

Some re-entering nurses were employed as regularly qualified and registered nurses, while other participants were recruited for more supporting roles, such as nurse assistant staffing and nurse aide staffing. Additionally, during the peak of COVID-19, a new position was implemented in hospitals, a so-called ‘buddy role’ to assist nursing staff in their daily tasks.

### Job description & task coordination

The majority of the participants argued that there was no clear job description allocated to their position. However, the extent to which division of tasks was coordinated differed per healthcare setting. Participants described working in rapidly established COVID-19 departments as pioneering and mentioned that a structured working method still had to be formulated. In practice, most declared to have equivalent jobs, performing every task within their ability and complementing each other.

> *“We were kind of told. ‘do whatever you can.’ You have to compare it to development aid. ‘Do what you can do, and if you have any questions, we will hear from you.’ With that message they kind of let us go.“ (Participant at COVID-19 department in a nursing home setting) R5*

Those working in nursing homes or at a rehabilitation centre mentioned that the interpretation of their role was often not discussed and that the interpretation mostly depended on their own perception. These re-entering nurses said they felt comfortable in their role due to the limited complexity of the care that they provided and its supportive nature.

> *“People on the ward knew that I was coming, but my role was not clear, it had not been communicated. It was a bit difficult because I was not sure myself either. I just introduced myself a bit, and they left it up to me to decide what I did and did not want to do.” (Participant at Nursing home) R6*

Re-entering nurses in a buddy role in a hospital setting were generally positive about the buddy system, which allowed them to coordinate the division of tasks together with a more experienced nurse. This position was often described as very task-oriented. Some re-entering nurses declared to perform specific chores delegated by nursing staff, such as washing patients, preparing medicine or stocking the department. Others mentioned independently taking care of a single patient per shift whilst nursing staff retained the ultimate responsibility and guided buddies by maintaining a helicopter view. The hospital-based participants working as independent nurses declared not to experience any unclarity with the scope of their role:

> *“At 7:30, you were linked to an ICU nurse, who told you what patients you were going to take care of. They said, ‘the idea is that everything I ask you to do, you essentially should do.’” (Participant buddy at ICU) R11*

Overall, the scope of practice for returners seems to be strongly linked to the extent to which re-entering nurses felt comfortable and proficient at executing the tasks aligned to their position. The importance of protecting one’s boundaries was commonly reported by participants. For example, most re-entering nurses in COVID-19 departments were employed as a nurse during their return but declared not feeling comfortable with the responsibility, thus took a more supporting role instead. Contrarily, a few nurses who returned to the ICU started as buddies, but mentioned to quickly get accustomed to nursing again and therefore stepped into the role of an independent nurse. However, this led to unusual situations. One participant who had not worked as a nurse for eight years mentioned:

> *“I was always scheduled as a buddy. However, they (referring to nursing staff) quickly realised that they could deploy me as an independent nurse, which is obviously madness because if you think about it…on my fifth day, I was taking care of my own patient. I only had been walking around there for five days, so that is kind of… it needs to be in your nature to dare to take on that responsibility. I did that with limited knowledge. I had my own buddy because of that, and that helped.” (Participant independent nurse at ICU) R7*

Even though re-entering nurses mostly felt comfortable in their role, in all settings participants mentioned examples of unclarity that was caused by the lack of a clear job description. Some participants said that expectations within teams deviated. For instance, one participant mentioned discovering that she was allocated to be the nurse with the final responsibility, while this had not been communicated to her. Another participant said that her new colleagues did not expect to get a re-entering nurse in their team:

> *“Well, they put me in a protective suit, gave me a mask and gloves and then I was allowed to go into the ward. There were already two people working, and I asked with whom I could tag along. Their reaction was: ‘tag along? Surely you can wash people?’ It was a bit like: ‘we cannot give instructions to people right now; we need a pair of extra hands on the bed.’” (Participant at COVID-19 department in a nursing home setting) R10*

Moreover, some participants mentioned that it was unclear what the limitations of their role were and what nursing skills they were authorised to execute or not. The authorisation concerning the execution of practical nursing skills or the reserved procedures (as described in the BIG-law) were handled differently per organisation. In some cases, re-entering nurses said that they agree with colleagues that they could not perform any reserved nursing procedures or high-risk practical nursing skills. Others declared to be allowed to perform these specific tasks when colleagues could confirm their competences or if they felt comfortable and proficient at doing so.

> *“Well, in the beginning I mentioned the BIG registration. Yes, I think it’s good that it exists -the BIG registration - but then the question is how flexible should we deal with this? Do you say someone is going to work again, and they will acquire their BIG registration again during that year, or do you say you can’t work until you are BIG registered again, that’s a bit*

… In this crisis situation, no one asked ‘gosh (participant’s name), please hand over your BIG registration.’ “ Participant at ICU) R3

Additionally, within nursing homes, some re-entering nurses mentioned confusion about the distinction in nursing skills. Not all practical nursing skills are reserved procedures, and some participants mentioned to question if healthcare staff was aware of the difference between practical nursing skills and the reserved nursing procedures:

> *“I believe that if you are not articulate enough and you get the confidence that you can do this, you might be tempted to do something for which you are not authorised. For me, that was with the replacement of catheter bags. Yes, I did not know myself either what I was allowed to do or not with that. Is that a reserved nursing procedure, or is it not? If that is unclear for them (referring to colleagues), then it is unclear for me, and there was, actually no one who told me at the beginning what I could or could not do.” (Participant at Nursing home) R2*

In light of the COVID-19 pandemic and its unexpected magnitude, most participants said that it was understandable that no clear task description was available. Re-entering nurses mentioned being flexible and open-minded towards their role. However, participants also said that a framework with a job description would have helped to clarify their position for themselves and their colleagues:

> *“It would have been nice for me personally, and the rest of the team, if there was an instruction sheet stating specific tasks I can do, and the ones I can leave for someone else to do. I think that would have helped a lot.” (Participant at a nursing home setting) R6*

### Supervision & guidance

The extent to which supervision and guidance for re-entering nurses was provided differed per healthcare organisation. Overall participants mentioned taking responsibility upon themselves to be proactive and ask questions to their colleagues. The re-entering nurses placed in a buddy role were linked to one particular nurse who maintained a helicopter view of the activities, and most buddies said that this provided adequate guidance. However, some of the re-entering nurses in the other settings who were not linked to healthcare staff mentioned that they would have preferred to have one specific person to whom they could turn to for advice and evaluation.

> *“In practice, I believe that it would be very good if someone would be linked to a specific mentor. I do not have that right now. For instance, tomorrow I have to work again, and then I look who is there and some I probably already know, and yes then you know how or what, but for re-entering nurses, I believe it is incredibly important to have a specific person to whom you could go with questions and other struggles.” (re-entering nurse in rehabilitation centre) R1*

### Training

The extent to which the re-entering nurses had received training varied greatly amongst all participants in all settings. Three re-entering nurses working in hospital settings mentioned to have received training in a skills lab and the opportunity to do e-learnings. All three participants reported being content with their received training. Contrarily, seven participants declared to have not received any kind of training prior to their return, and the majority of the other participants said that they only had the opportunity to perform (mostly non-obligatory) e-learnings. Some participants mentioned getting a short orientation of the department, instructions on how to wear PPE and how to turn COVID-19 patients in the prone position. Overall participants said that it was unrealistic to wish for a complete training prior to their return, considering the circumstances of the pandemic.

> *“There was no time to do so; there really was not. Everyone was in an uproar and the nurses who could have done this were all working, so it was take-it-or-leave-it”. (Participant buddy at ICU) R11*

A few participants mentioned faring well in their position despite the lack of training. Participants in supporting roles often mentioned being still knowledgeable in basic nursing actions, such as providing personal care, for example, assisting in washing, getting dressed and eating and communication.

> *“I did not experience that as something I missed. Absolutely not, because it was just like riding a bicycle; basic care is something that you do not forget, I could do that.” (Participant at nursing home) R6*

### Training content

Nevertheless, when participants were asked about an ideal returners programme to be prepared for their return into nursing, some general wishes did derive. Most returners said that a theoretical training programme was unnecessary, yet mentioned the need to refresh their practical nursing skills. Furthermore, participants mentioned missing an orientation concerning the basic working methods of the departments, such as the daily routines and electronic patient records. Another common view amongst participants is that training must align with the individual level of the returner and should be based upon specific skills or knowledge that need some repetition. Many re-entering nurses mentioned that returners should not be treated as new students, whereas they carry their own life experiences and have obtained other useful skills over time.

> *“Returners are people who often have quite some life-experience. It would be nice if training relates to this, so they do not have to do everything by default, but where you look at what someone needs. I have been out of the running for 28 years, but someone who was only absent for five years needs entirely something else.” (Participant at nursing home) R1*

Additionally, some needs concerning training varied per healthcare setting. Several re-entering nurses at ICUs mentioned encountering situations in which they struggled with treatments, protocols and medical equipment such as ventilators and monitors that had been changed. Meanwhile, those who re-entered in a nursing home setting mentioned encountering innovative patient-lifts systems and electronic devices such as thermometers which they were unfamiliar with.

> *“I think that for the technical procedures, they should have included a moment in the beginning to show the devices. For example, the thermometer that you hold against your head, you know? Yes that is for example something that I had not done before and I did not know.” (Participant at COVID-19 department within nursing home setting) R9*

### Positive team dynamic

The majority of the re-entering nurses said that there was a positive team dynamic within the departments as a consequence of the COVID-19 pandemic. The recruitment of many different (former) health professionals from a variety of backgrounds led to diverse and sometimes new teams. Some participants explained that it was challenging to get acquainted with each other as a team, due to the hectic pace of the COVID-19 pandemic and covering PPE. However, despite the variety within teams, participants often mentioned experiencing strong commitment and solidarity within teams to counter the challenges of the pandemic collectively. Several re-entering nurses said that the pressure to combat a novel emerging disease resulted in a shared vision and the need to collectively focus towards shared results, which resulted in positive team dynamics.

> *“Something that I really liked was that you are part of the team right away, the solidarity was there from day one, so that is how I experienced it. And you can see that everyone is trying their best, and it might be a little stressful, but it is all from a good heart. So that is beautiful to see and that you are part of that is very cool that you do it together.” (Participant at a COVID-19 department within a nursing home setting) R10*

One participant did mention that at certain moments the buddy system led to situations in which he felt less included. According to this participant, the current system sometimes created a pecking order amongst buddies.

> “For example, a colleague said, ‘I am fine with taking care of that patient, but then I do want a good buddy.’ The room was filled with all the buddies, so when they picked someone, you knew that was a good buddy, and the rest was rubbish. You have to imagine that you are completely dressed up with a mask and eye protection, and then that is being said. I could not comprehend it.” (Participant buddy at ICU) R7

### Mental health

Several nurses who directly worked with COVID-19 patients did experience the pandemic as an intense and sometimes emotional period. Participants explained that the unpredictable clinical picture of COVID-19 and the confrontation with the passing of patients was overwhelming. Additionally, the longer working hours, extra shifts, and the uncertainty of the course of the pandemic was described as demanding by some re-entering nurses. Few participants mentioned fearing contamination with COVID-19 and said to be especially concerned for their family. In these cases working in protective equipment and continuously being aware of possible cross-contamination was experienced as stressful and demanding.

> *“At that moment, it did not really hit me, but then I got back, took a shower, and I wanted to go to sleep, but I could not fall asleep; I was restless. The next day my husband was going for a walk with my daughter, and I was home alone. I took the newspaper, and I saw an obituary with such a beautiful poem. I read the poem, and I started to bawl, like bawling and I bawled my eyes out, all emotions came out. That obituary triggered me, and that is when it hit me. That paper with five pages of obituaries, all from villages in (name of a province). Yes, that did something to me.” (Participant at COVID-19 department in nursing home setting) R5*

However, despite these findings, nearly all participants mentioned that the re-entering process did not lead to the impairment of their mental health. In contrast, some participants even mentioned that their re-entry had improved their mental health. Re-entering nurses said that they enjoyed their role, the contact with patients and were happy to contribute in a time of need. Moreover, participants mentioned feeling appreciated. The nursing departments received words of support, free food, care packages and even massage chairs. Additionally, some participants mentioned experiencing less pressure due to responsibility because of the supporting nature of their role.

> *“It really did not affect me negatively. We were all just very combative, we all went for it, and that has all just been very positive. No, I did not burn-out or became very stressed because of this. On the contrary, I thought it was a cool period actually.” (Respondent buddy at ICU) R11, Q17*

### Mental health support

Even though mental health support was facilitated in the form of counselling by psychologists for those of whom worked with COVID-19 patients, nearly all participants explained that they did not feel the need to use this possibility. Most participants mentioned being able to cope with their mental health independently. Some returners mentioned discussing their experiences with family members, while others declared to frequently sit together with colleagues to evaluate and support each other.

> *“ If we were wearing those protecting suits, then we could hold on to each other. Yes, outside of course we could not, but on the ward, we had those protecting suits and then we would hold on to each other for a bit. The beautiful thing was that even though no one knew each other in this department, we could shed a tear together. That gave a wonderful feeling.” (Participant at COVID-19 department within nursing home setting) R5, Q18*

## Discussion

This qualitative study investigated experiences and needs of former nurses who re-entered during the first wave of the COVID-19 pandemic in the Netherlands.

We showed that a lack of a clear job description with operationalised roles, responsibilities and restrictions sometimes led to unclarity about role division and the scope of practice and deviating expectations amongst returners and healthcare staff. This unclarity was especially notable in the newly established COVID-19 departments. Participants mentioned that due to the rapid establishment of the departments and the new teams, a structured working mechanism was not yet established. In all settings, re-entering nurses said that the fulfilment of roles was closely related to the extent to which they felt comfortable and proficient, and participants emphasised the importance of protecting their boundaries. Subsequently, participants mentioned that they often decided to take a more supportive position. However, in some cases, the uncertainty on occupational boundaries led to re-entering nurses executing reserved procedures that are normally considered to be reserved for up-to-date educated registered nurses. This is in line with the findings of Liberati (2017) and Xyrichis (2017) who observed an increase in informal crossing of boundaries due to work urgency, such as during a pandemic. Even though this does not necessarily have to lead to the impairment of quality of healthcare, it is a risk that blurred demarcation between roles leads to the informal shifting of tasks beyond the scope of practice of lay healthcare workers (Callaghan et al., 2010). In the same light, historical research on the influenza pandemic of 1918-1919 in New-Zealand showed that nursing staff perceived vague boundaries between lay nurses who were employed during the pandemic and the professional nursing staff as a threat to the quality of care (Wood, 2017). A transparent description of roles and the associated competencies is essential to create clarity on one’s responsibilities and accountability (World Health Organisation, 2008; Munga et al., 2012; Zachariah et al., 2009; Callaghan et al., 2010; ledikwe et al., 2013).

The results of this study further indicated the usefulness of an easily accessible mentorship structure for re-entering nurses based on a collaborative approach between, for instance, experienced nurses and those who re-entered. We found that former nurses in a buddy role who were linked to other nurses were especially content about the accessibility of guidance. Supervision is a well-known essential factor for the retainment of nurses and having the opportunity to consult with a mentor increases confidence and competences (Durand & Randhawa, 2002; Long & West, 2007; Mark & Gupta, 2002; Myall et al., 2008; Pellatt, 2013). Moreover, research on nursing at a COVID-19 ICU with an implemented buddy system reported the approachable mentorship of experienced ICU nurses to be a valuable model to guide nurses who are normally not employed in ICU settings (Marks et al., 2020).

Further, our findings indicate the need for an individualised and mainly practical training program for re-entering nurses, harmonised with their current level of competences and knowledge disparities to become skilled enough to assist in nursing (during a pandemic). The majority of the participants had received a limited amount of training to master complex nursing procedures due to their rapid return, which seemed partly dependent on the healthcare setting where they re-entered. Re-entering nurses come from different backgrounds, therefore obtain different levels of experience in nursing. Yet, most re-entering nurses mentioned that they were still able to perform basic nursing care, such as supporting patients with personal hygiene, clothing and eating, despite the numerous years of not practising. In line with the findings of this study, literature shows that re-entering nurses do not want to be approached as new nurses; hence like to be appreciated for the skills they already possess (Barriball et al., 2007).

The situation of the pandemic led to a shared vision and purposeful collaborative teamwork which created positive team dynamics, despite the divergent backgrounds of the team members. Participants mentioned that the pressure to combat a novel emerging disease resulted in the need to focus towards shared results collectively. This finding was also identified in previous research on nursing during healthcare crises, in which measures of appreciation, support and feelings of unity led to a high working moral (Renke et al., 2020; Corley et al., 2010; Biai, 2007). Moreover, Shih et al., (2002) showed that sharing the experience of providing care after the earthquake of 1999 in Taiwan and the urgent need to help each other, led to a caring relationship amongst team members.

The participants said that the negative impact of returning during the COVID-19 pandemic on their mental health was limited. In line with literature, some former nurses who were exposed to infected patients experienced the pandemic as an overwhelming, emotional, uncertain and demanding period (Lai et al., 2020). However, contrary to expectations, nearly all participants in this study reported that re-entering in these circumstances did not lead to the impairment of mental health. Several factors might have influenced the limited impact of re-entering during a pandemic on the mental health of re-entering nurses. Firstly, most re-entering nurses had supporting roles during their re-entry; consequently, some participants said that they experienced less pressure of responsibility. Furthermore, many participants mentioned experiencing solidarity within teams and strong team coherence. Literature shows that a sense of coherence and social support in the workplace could function as a protecting factor for one’s mental health (Greenberg et al., 2020.; Malinauskien et al., 2009; Wats et al., 2013). Moreover, most re-entering nurses mentioned feeling appreciated. During the pandemic nurses were supported by kind words and gestures of food and gifts by their environment. Research shows that a culture of appreciation in the workplace decreases the chances of burnout (Maslach & Leiter, 2017). Nevertheless, we should take into consideration the possible long-term psychological consequences of re-entering during a pandemic. Most re-entering nurses had not returned to practise for longer than two months at the time of the interview. It is questionable if prolonged exposure to working during a pandemic with multiple waves will increase the risk of impaired mental health. Research on COVID-19 and literature on previous pandemics show that a high number of healthcare workers experience severe mental health issues as a consequence of working on the frontlines of disease outbreaks (Khalid et al., 2016; Maunder et al., 2003; Pappa et al., 2020).

### Strengths and limitations

The main strength of this study is that the collection of data was executed during the COVID-19 pandemic, while participants were still working as a returner or had only just resigned, which decreased the chances of recall bias. Moreover, the topic is urgent and we provided important insights in the experiences of re-entering nurses.

For this study, we chose to include former nurses who re-entered in different healthcare settings. Due to the small number of re-entering nurses per healthcare setting, we were only able to identify main themes that generally apply to re-entering nurses during a pandemic, while full saturation on sub-themes for each specific setting was not achieved.

### Recommendations for future research

The perspective of current healthcare staff working with re-entering nurses is an essential topic for future research, considering the influences of re-entering of nurses during a pandemic on current healthcare staff. Mentoring, supervising and correctly assessing the proficiency of re-entering nurses during the already hectic circumstances of a pandemic, might be very straining on current healthcare staff.

## Conclusion

The rapid need for former nurses to return in times of the COVID-19 pandemic led to limited time to prepare and establish a structured re-entering process. These circumstances often cause uncertainty about roles and responsibilities amongst re-entering nurses. This was especially challenging in the newly established COVID-19 wards within nursing home settings, while the process appeared less uncertain and chaotic within highly-structured organisations, such as at ICUs in hospitals. However, despite the challenges of the re-entering process, the re-entering nurses maintained an open-minded and flexible attitude. The situation of the pandemic led to purposeful collaborative teamwork, and re-entering nurses generally did not report negative impact on their mental health. The results of this study indicate that the following is needed to support a rapid and safe return: a clear description of roles and responsibilities; an individualised assessment determining the competences and knowledge disparities of re-entering nurses; practical training focussing on competencies needed during a pandemic; and a responsive mentorship structure to guide re-entering nurses.

### Relevance to clinical practise

In light of the current ongoing COVID-19 pandemic and possible emerging new pandemics in the future, it is essential to consider strategies to rapidly increase health care capacity. The rapid recruitment of former nurses to mitigate an acute shortage of qualified nurses could play a vital role during the current and future pandemics. In order to ensure the quality of healthcare, prevent problems and protect employability and resilience of re-entering nurses, this research provides insights into the re-entering process and addresses the needs of re-entering nurses during a pandemic.

## Data Availability

Transcripts are not available due to traceable information to individuals and institutions.

## Acknowledgements

The authors thank all re-entering nurses who participated in the interviews for participating in this study

## Funding or Sources of Support

This research did not receive any form of funding, nor sources of support

## Contributions

Study design: TH, MOK, SAN Data collection: SAN

Data analysis: TH, MOK, SAN Manuscript preparations: TH, MOK, SAN

## Appendix 1

Appendix one provides the conceptual background that was used for this research. While the COVID-19 pandemic provides a unique situation, we drew inspiration from existing literature on nursing during a pandemic, and re-entering nurses, which offered relevant insights for the development of the study protocol, an interview guide and data collection.

### Training

Former nurses re-entering during a pandemic need to rapidly update existing knowledge and competencies and develop new ones to become proficient in nursing during a pandemic. Research shows the need for former nurses to be aware of infection control principles and be agile in the use of personal protection equipment (PPE) (Chan & Wong, 2007; Irvin et al., 2008; Martin, 2011; McMullan et al., 2016). Nurses are the primary caregivers of vulnerable and susceptible patients and are in close contact with diseased patients. Therefore, nurses must have the knowledge to recognise and screen for possibly infected patients (Chan & Wong, 2007; Martin, 2011). Additionally, Research on re-entering nurses shows that former nurses prefer practically oriented and training focussing on competencies that are needed to work at a specific department (Durand & Randhawa, 2002; Long & West, 2007).

### Role division

Less qualified nurses rapidly re-enter care, which could create unclarity about the division of roles. Existing literature on task-shifting implies that unclarity on the division of roles often led to friction between healthcare workers; the hierarchy changed, since it became obscure who was responsible for what tasks (Callaghan et al., 2010). Zachariah et al. (2009) showed that not all healthcare workers felt comfortable with the additional supervisory responsibilities and the delegation of tasks due to task-shifting.

Additionally, unclarity on roles may lead to quality issues. Key-findings from an evaluating study on task-shifting in Botswana revealed tasks-shifting workers often performed more activities than they were educated and commissioned for (Ledikwe et al., 2013). A clear demarcation of responsibilities, tasks and boundaries including the advice of the involved health workers could substantiate the process of task-shifting (Callaghan et al., 2010; Ledikwe et al., 2013; Zachariah et al., (2009).

### Supervision & support

Former nurses who rapidly re-entered into care with limited preparation time to master complex nursing tasks and in times of a pandemic are in need of sufficient supervision Research on the H1N1 influenza pandemic showed that junior nurses who had to rapidly skill-up in order be able to provide a high number of patients of advanced therapy, experienced feelings of anxiousness and stress due to a lack of supervision (Corley et al., 2010). Moreover, Mark and Gupta (2002), mentioned the lack of supervision as one of the main challenges for re-entering nurses. Research has observed re-entering nurses often worried about their competences or felt as if they were thrown into the deep end (Durand & Randhawa, 2002; Mark & Gupta, 2002)

### Mental health needs

Several studies have postulated about the adverse psychological effects of nursing during the COVID-19 pandemic (Kang et al., 2020; Lai et al., 2020; Zhu & Xu et al., 2020). Research indicates that nurses responding to the COVID-19 outbreak in Wuhan China often experienced anxiety and stress, which was associated with the long working hours, extra work pressure, close contact with infected patients and the lack of PPE (Lai et al., 2020; Zhu & Xu et al., 2020). A key factor contributing to fear amongst nurses was spreading of the disease amongst colleagues and family members and the increasing number of deaths (Lai et al., 2020; Zhu & Xu et al., 2020).

### Private life

Providing care during a pandemic could have a demanding influence on the personal life of former nurses. Existing research on the preparedness for an influenza pandemic recognised worries amongst nurses about shortages in staff, leading to a demand to work extra shifts and longer hours (McMullan et al., 2016). Moreover, research showed that nurses worried about exposing their environment to an increased risk of infection (McMullan et al.,2016; Corley et al., 2010). In two additional analyses of the willingness to work during a pandemic in America, nurses mentioned similar worries (Irvin et al., 2008; Martin, 2011). As a solution, McMullan et al. (2016) noted the importance of available resources and information to reassure nurses and their families.

## Appendix 2

Appendix two contains the interview guide that was used during data collection.

The following questions were asked during the recruitment of participants to assure a varying sample of former nurses working in different settings.

What is your age?
What is your profession?
How many years of experience do you have as a nurse?
In which department(s) and in what kind of institution(s) (hospital, nursing home, home care etc.) did you work?
Did you follow any nursing specialisations? If so: what is your nursing specialty?
In what year did you stop working as a nurse?
Were you BIG registered at the time of your re-entry into nursing?
In which department and in what kind of institution are you working momentarily as a nurse?
How long have you been back in nursing practise again?

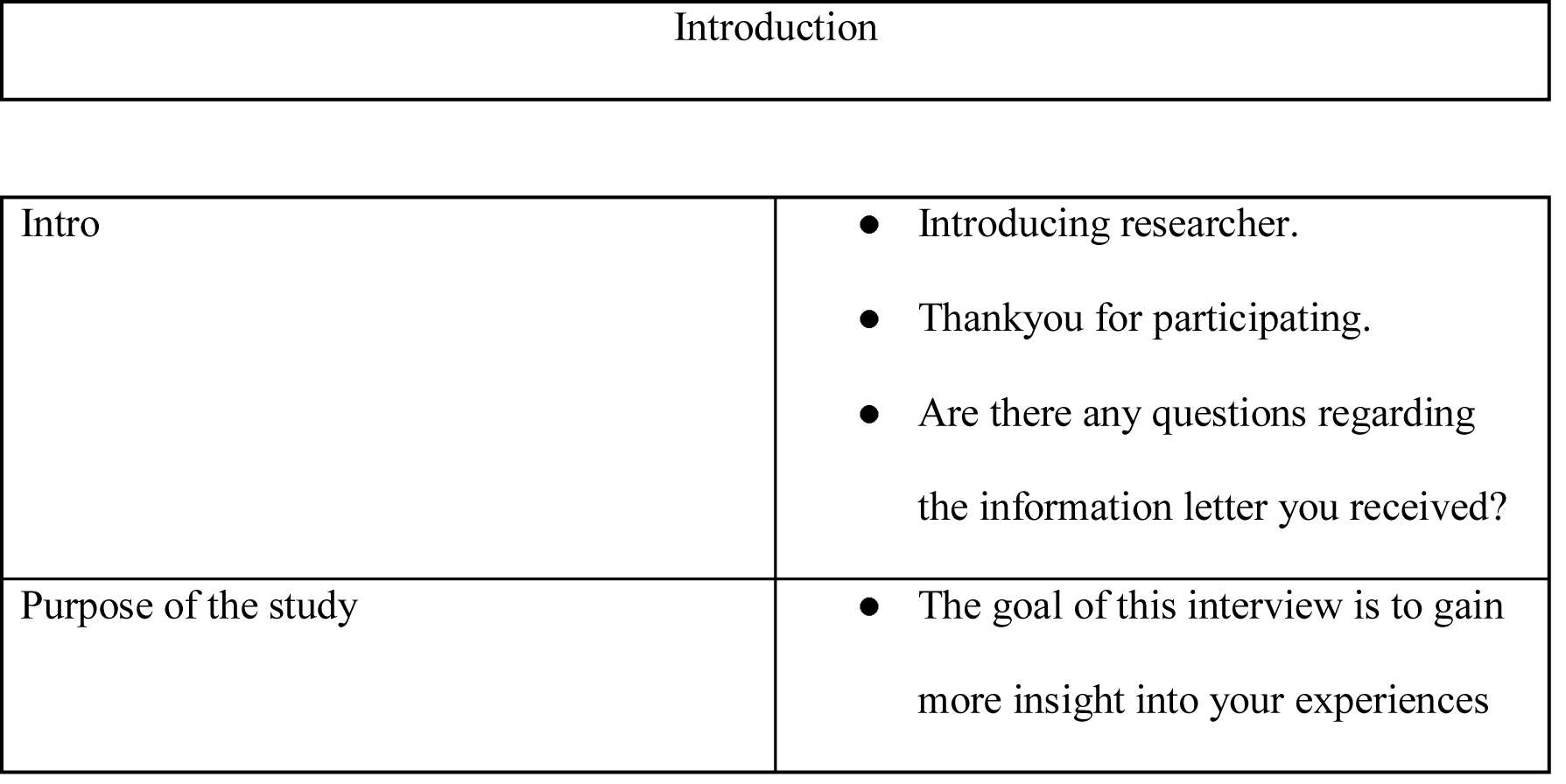
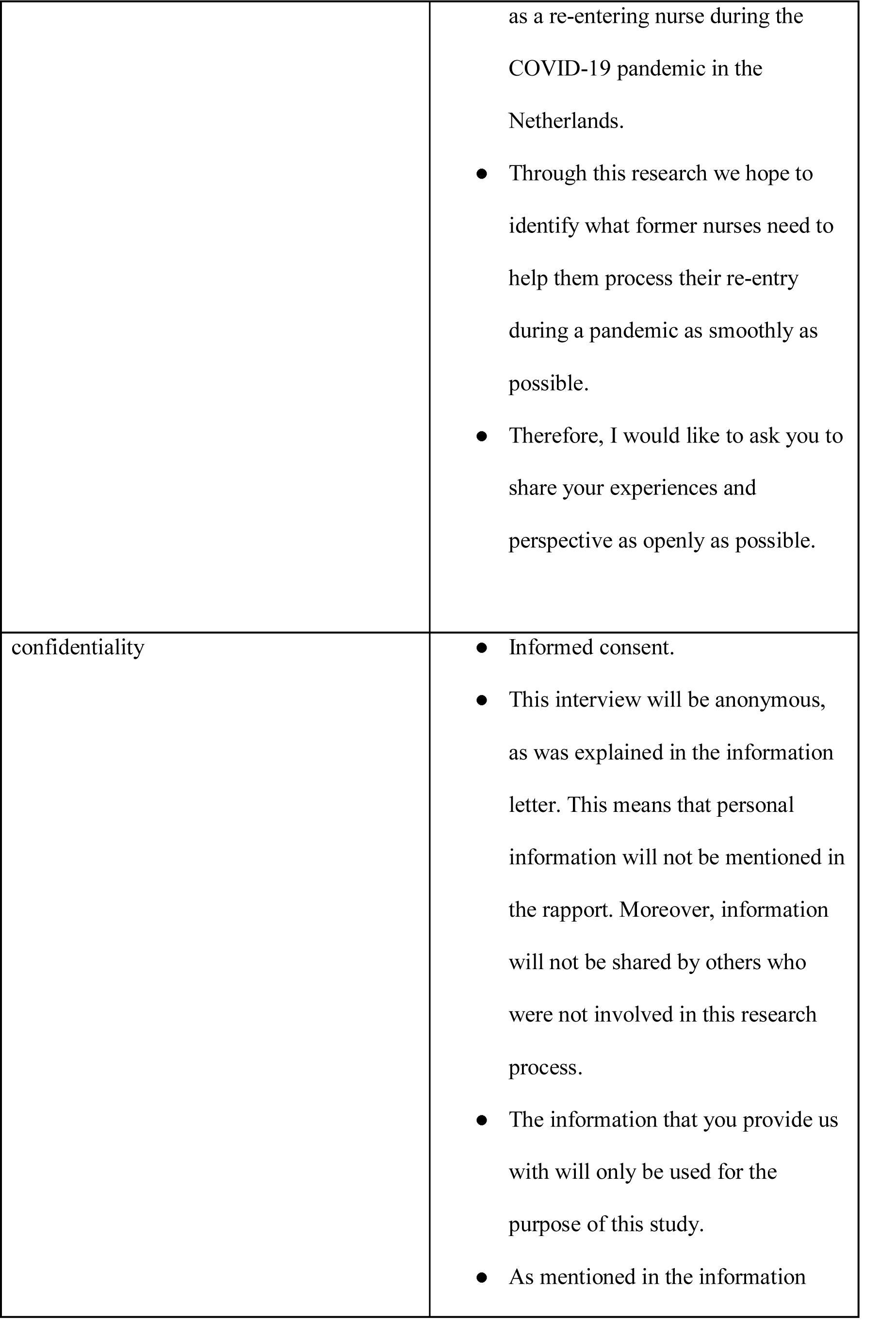
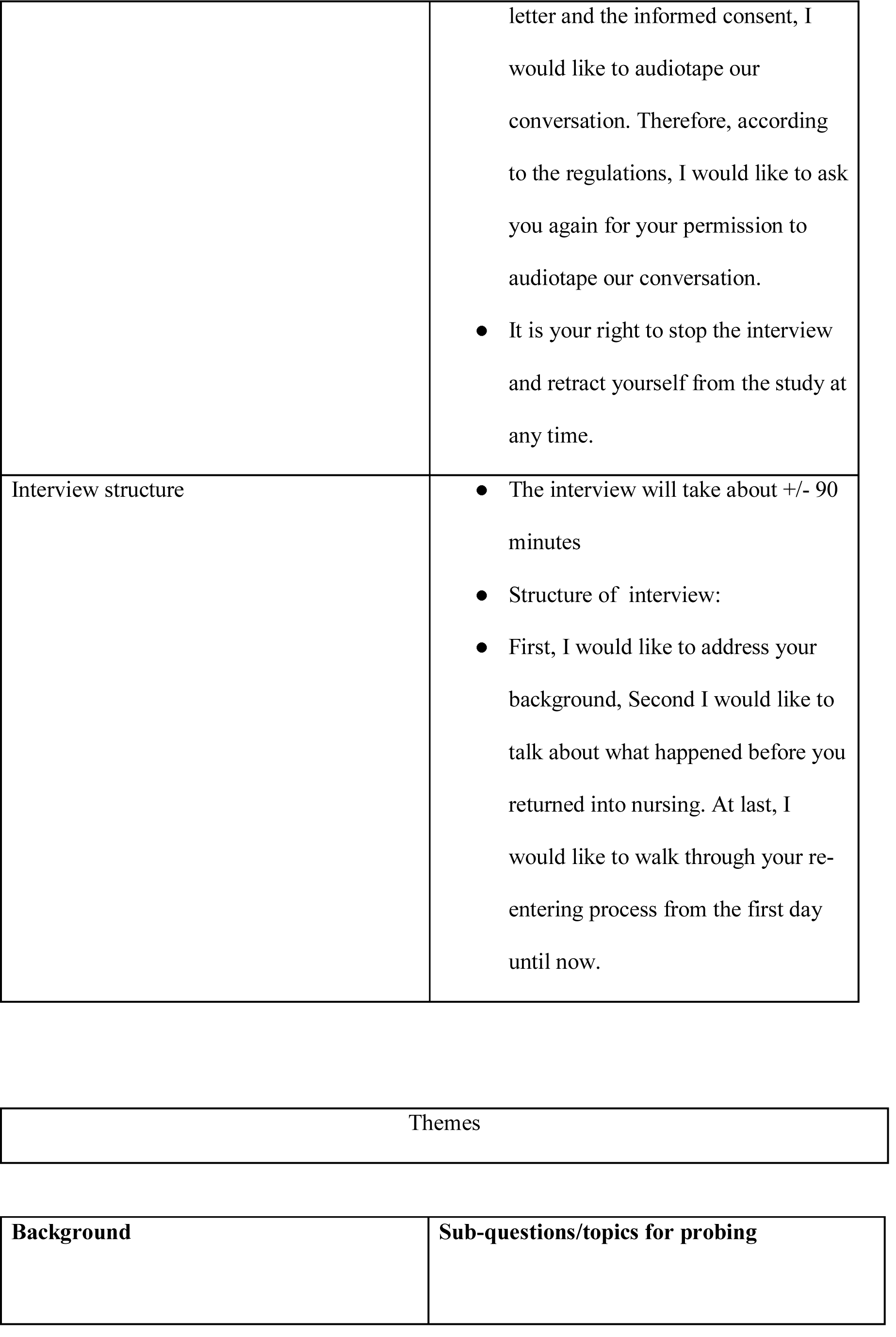
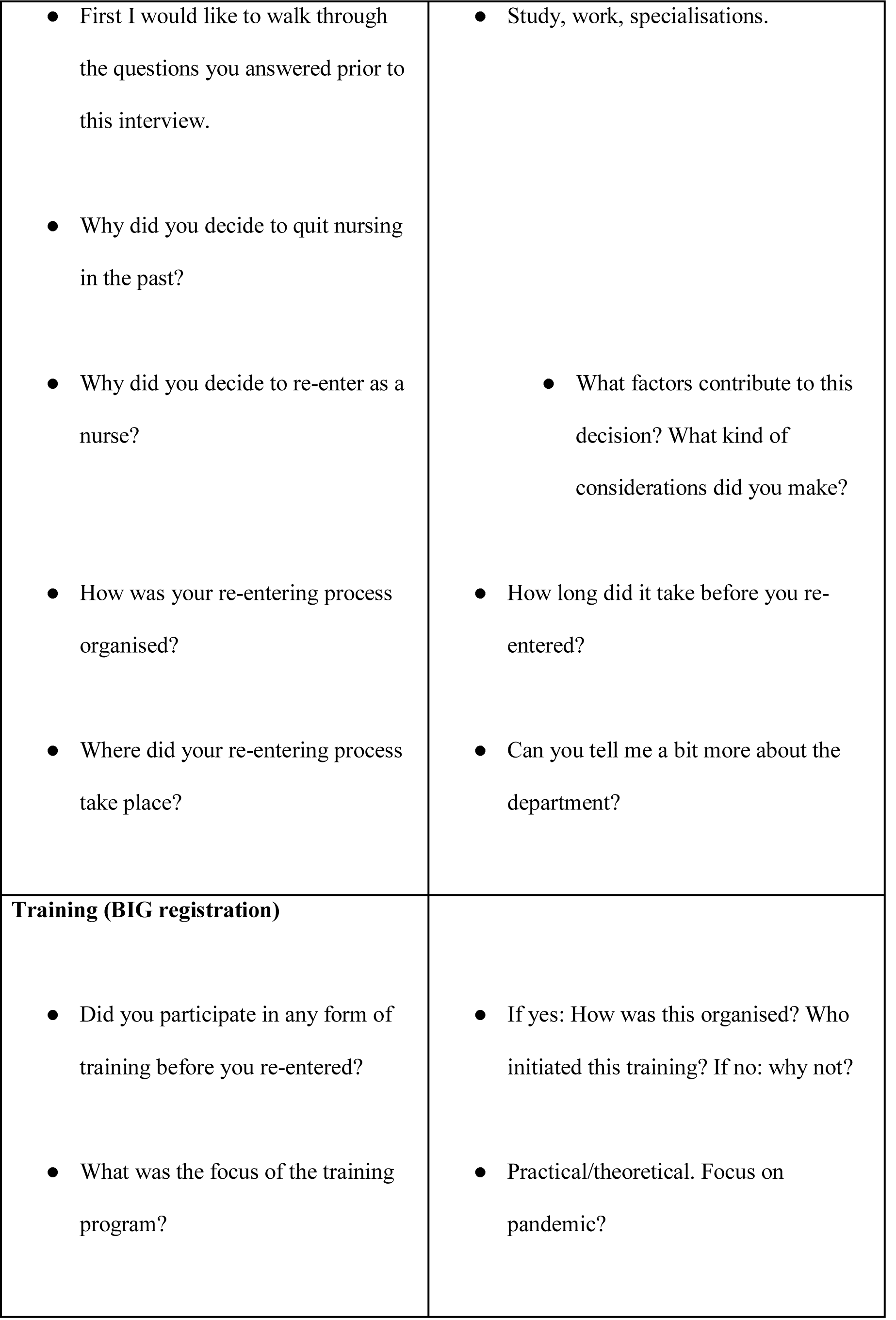
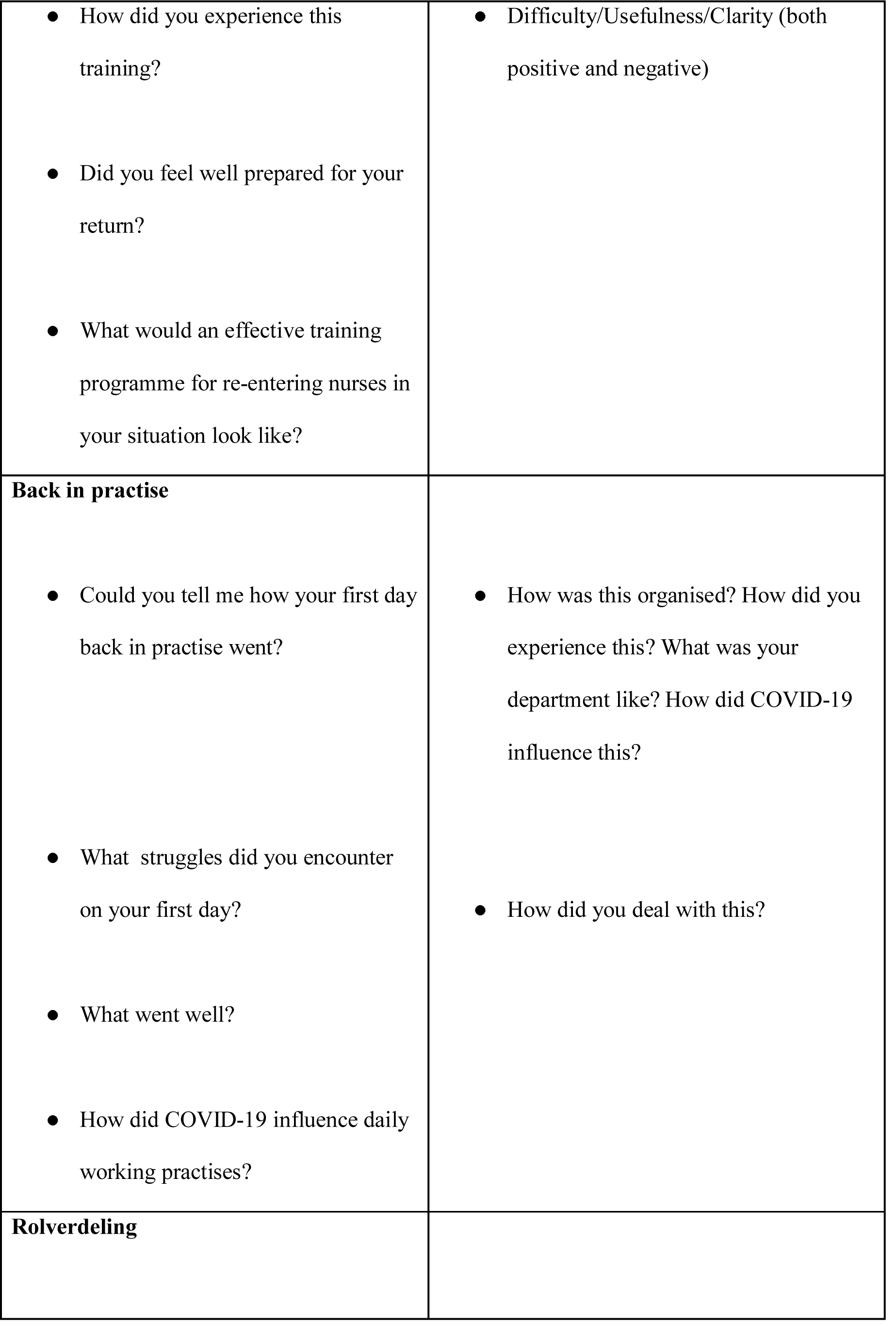
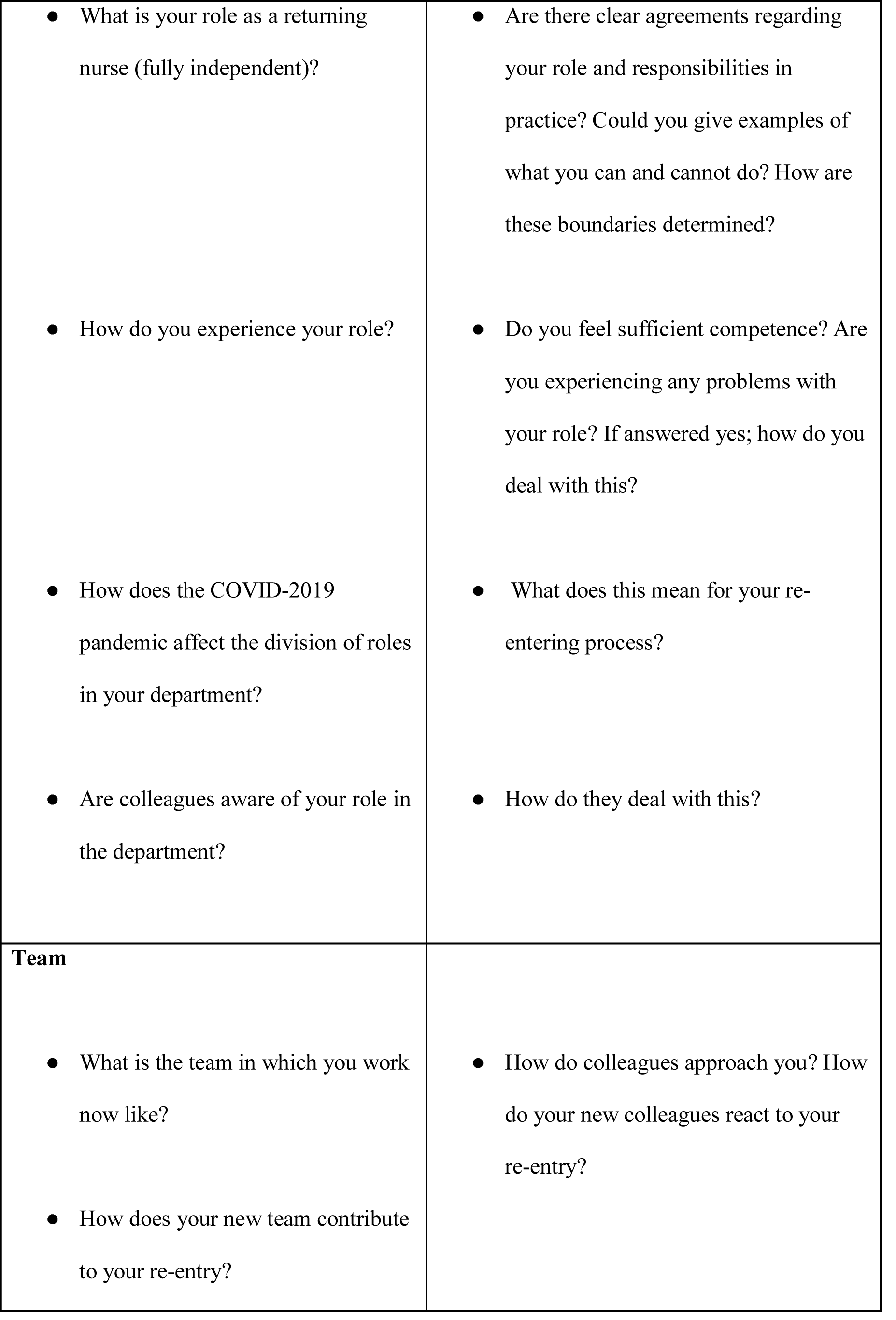
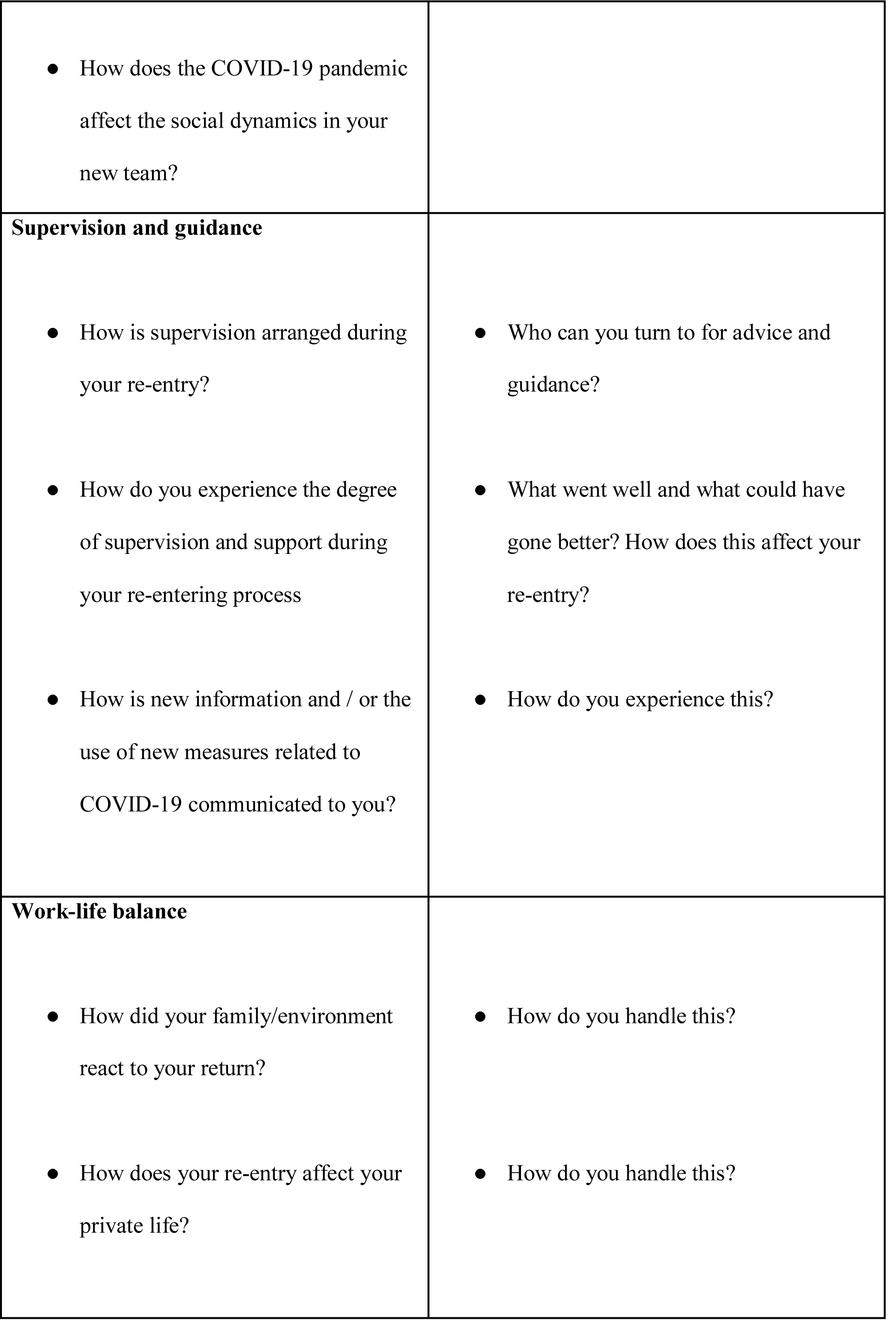
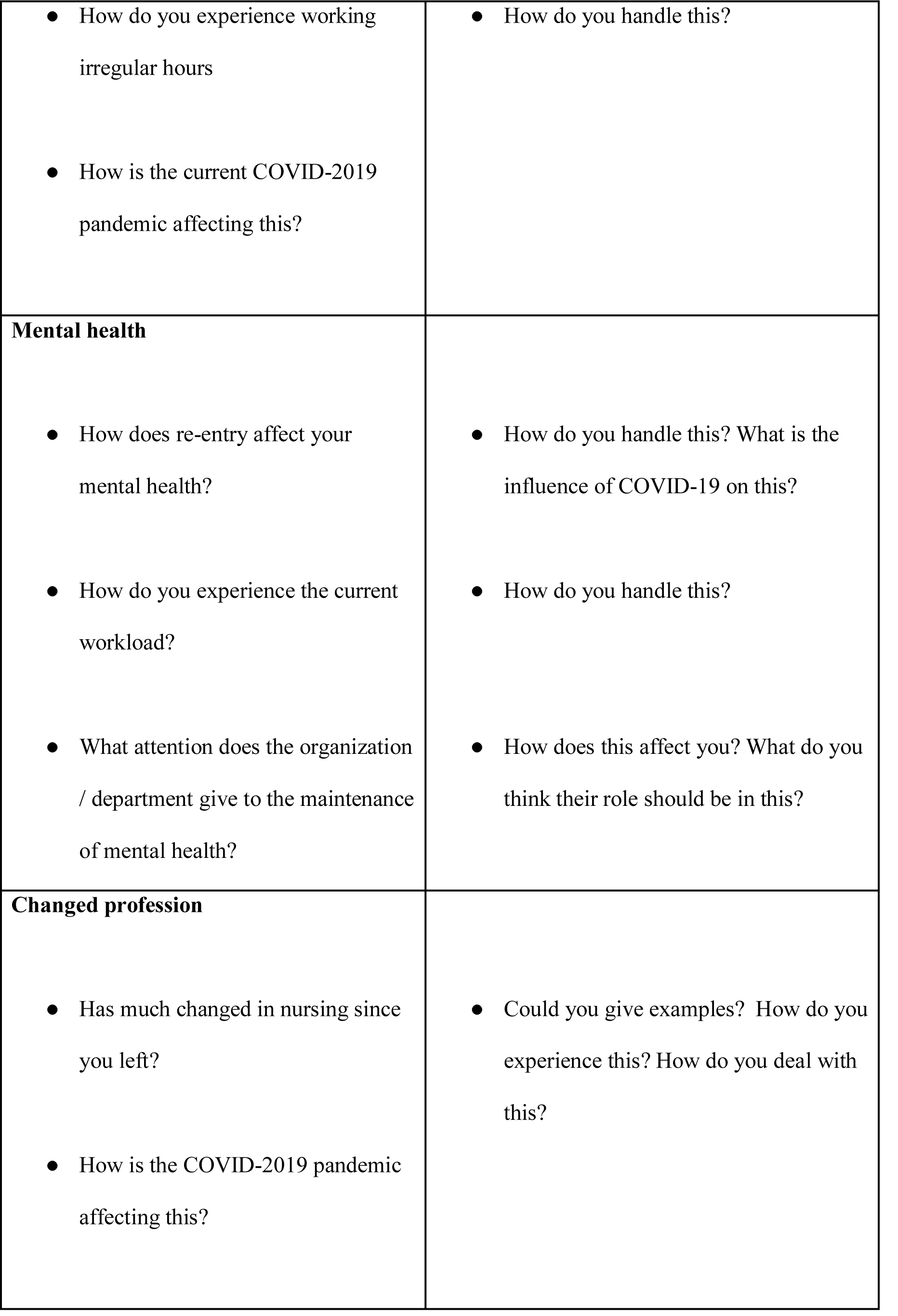
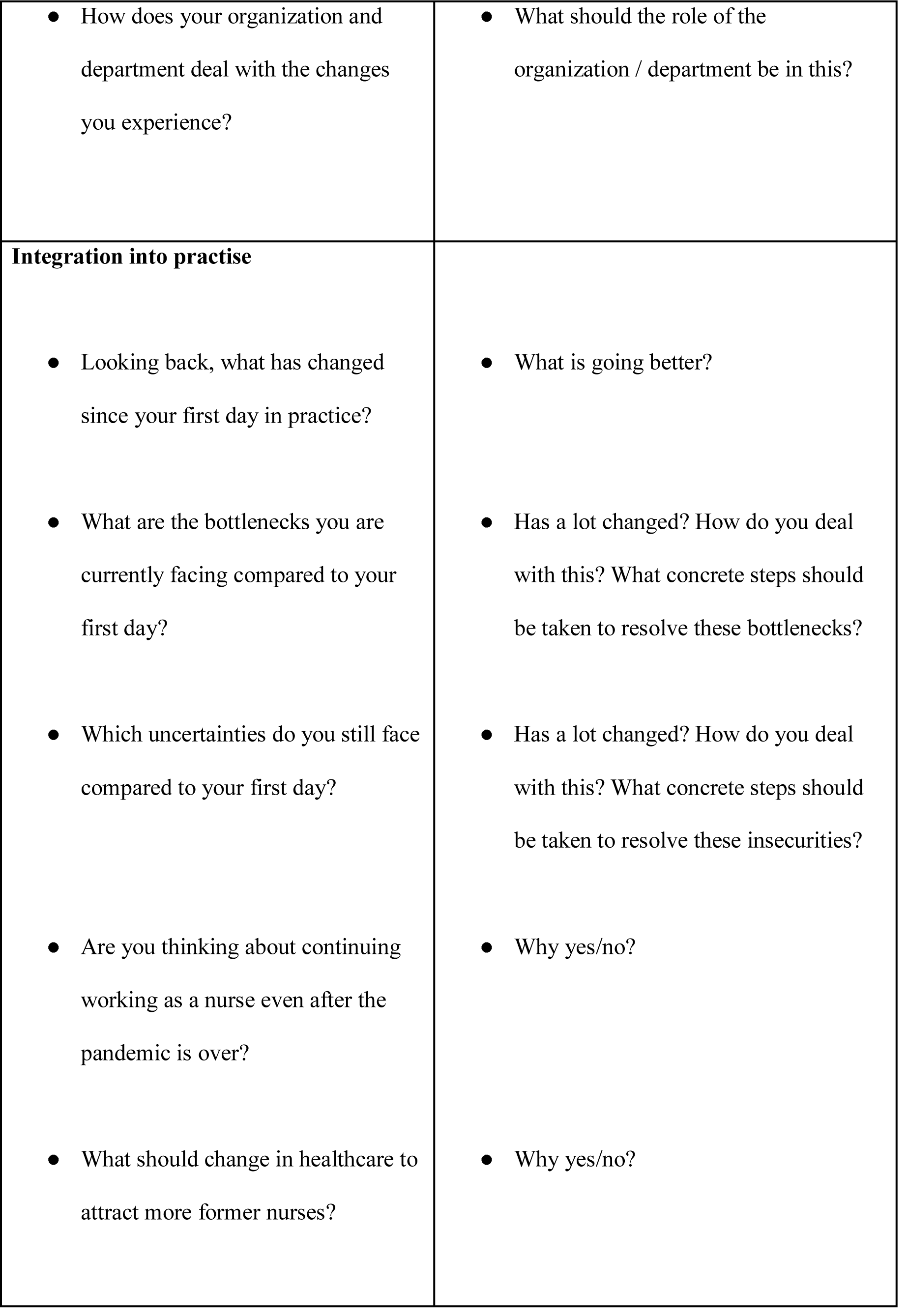
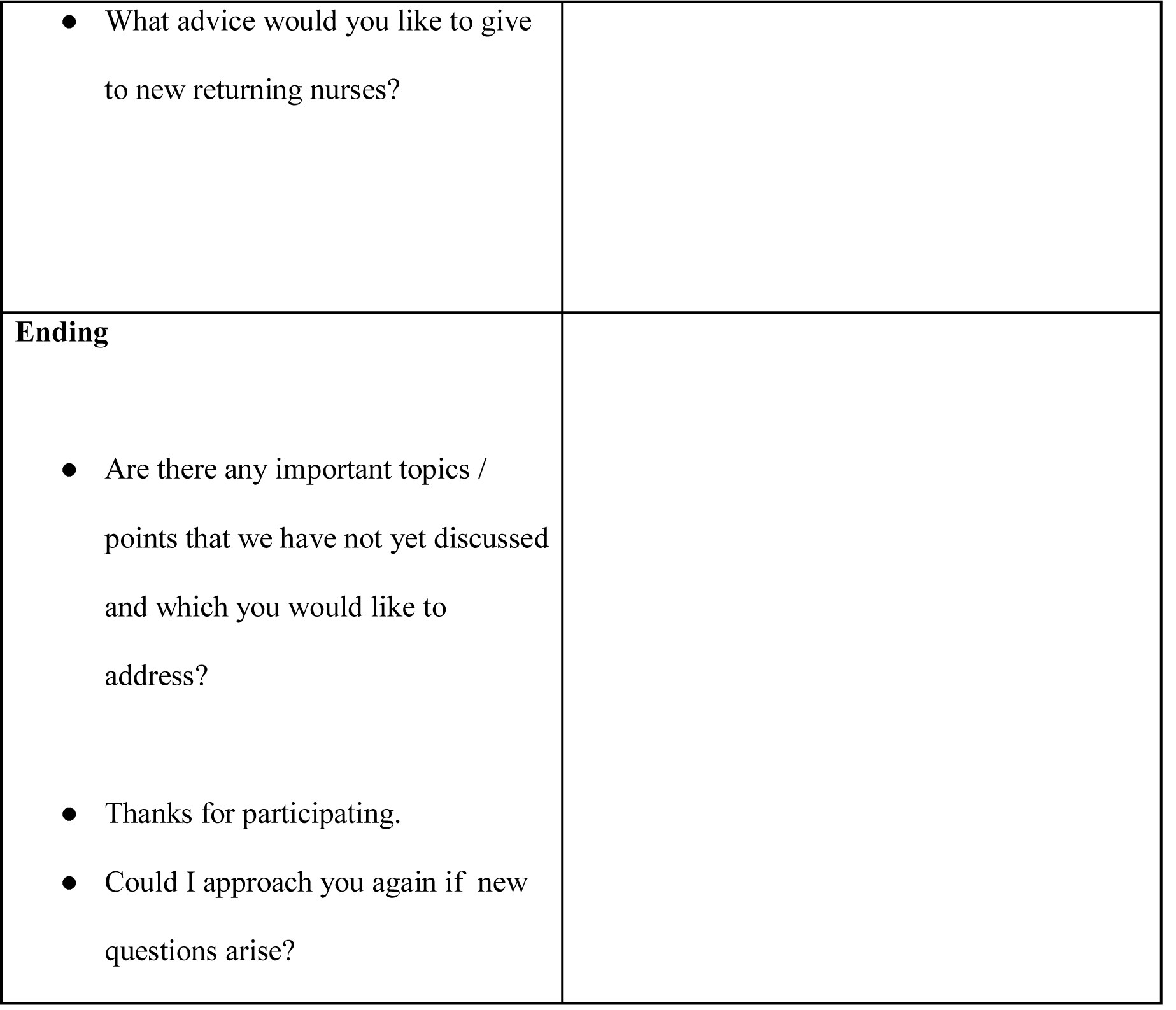

|  |
| --- |
| Introduction |

|  |  |
| --- | --- |
| Intro | ● Introducing researcher.<br>● Thankyou for participating.<br>● Are there any questions regarding the information letter you received? |
| Purpose of the study | ● The goal of this interview is to gain more insight into your experiences |
|  | as a re-entering nurse during the COVID-19 pandemic in the Netherlands.<br>• Through this research we hope to identify what former nurses need to help them process their re-entry during a pandemic as smoothly as possible.<br>• Therefore, I would like to ask you to share your experiences and perspective as openly as possible. |
| confidentiality | • Informed consent.<br>• This interview will be anonymous, as was explained in the information letter. This means that personal information will not be mentioned in the rapport. Moreover, information will not be shared by others who were not involved in this research process.<br>• The information that you provide us with will only be used for the purpose of this study.<br>• As mentioned in the information |
|  | letter and the informed consent, I would like to audiotape our conversation. Therefore, according to the regulations, I would like to ask you again for your permission to audiotape our conversation.<br>● It is your right to stop the interview and retract yourself from the study at any time. |
| Interview structure | ● The interview will take about +/- 90 minutes<br>● Structure of interview:<br>● First, I would like to address your background, Second I would like to talk about what happened before you returned into nursing. At last, I would like to walk through your re-entering process from the first day until now. |
| Themes |

|  |  |
| --- | --- |
| **Background** | **Sub-questions/topics for probing** |
| • First I would like to walk through the questions you answered prior to this interview.<br>• Why did you decide to quit nursing in the past?<br>• Why did you decide to re-enter as a nurse?<br>• How was your re-entering process organised?<br>• Where did your re-entering process take place? | • Study, work, specialisations.<br>• What factors contribute to this decision? What kind of considerations did you make?<br>• How long did it take before you re-entered?<br>• Can you tell me a bit more about the department? |
| **Training (BIG registration)**<br>• Did you participate in any form of training before you re-entered?<br>• What was the focus of the training program? | • If yes: How was this organised? Who initiated this training? If no: why not?<br>• Practical/theoretical. Focus on pandemic? |
| • How did you experience this training?<br>• Did you feel well prepared for your return?<br>• What would an effective training programme for re-entering nurses in your situation look like? | • Difficulty/Usefulness/Clarity (both positive and negative) |
| **Back in practise**<br>• Could you tell me how your first day back in practise went?<br>• What struggles did you encounter on your first day?<br>• What went well?<br>• How did COVID-19 influence daily working practises? | • How was this organised? How did you experience this? What was your department like? How did COVID-19 influence this?<br>• How did you deal with this? |
| **Rolverdeling** |  |
| • What is your role as a returning nurse (fully independent)?<br>• How do you experience your role?<br>• How does the COVID-2019 pandemic affect the division of roles in your department?<br>• Are colleagues aware of your role in the department? | • Are there clear agreements regarding your role and responsibilities in practice? Could you give examples of what you can and cannot do? How are these boundaries determined?<br>• Do you feel sufficient competence? Are you experiencing any problems with your role? If answered yes; how do you deal with this?<br>• What does this mean for your re-entering process?<br>• How do they deal with this? |
| **Team**<br>• What is the team in which you work now like?<br>• How does your new team contribute to your re-entry? | • How do colleagues approach you? How do your new colleagues react to your re-entry? |
| • How does the COVID-19 pandemic affect the social dynamics in your new team? |  |
| **Supervision and guidance**<br>• How is supervision arranged during your re-entry?<br>• How do you experience the degree of supervision and support during your re-entering process<br>• How is new information and / or the use of new measures related to COVID-19 communicated to you? | • Who can you turn to for advice and guidance?<br>• What went well and what could have gone better? How does this affect your re-entry?<br>• How do you experience this? |
| **Work-life balance**<br>• How did your family/environment react to your return?<br>• How does your re-entry affect your private life? | • How do you handle this?<br>• How do you handle this? |
| • How do you experience working irregular hours<br>• How is the current COVID-2019 pandemic affecting this? | • How do you handle this? |
| **Mental health**<br>• How does re-entry affect your mental health?<br>• How do you experience the current workload?<br>• What attention does the organization / department give to the maintenance of mental health? | • How do you handle this? What is the influence of COVID-19 on this?<br>• How do you handle this?<br>• How does this affect you? What do you think their role should be in this? |
| **Changed profession**<br>• Has much changed in nursing since you left?<br>• How is the COVID-2019 pandemic affecting this? | • Could you give examples? How do you experience this? How do you deal with this? |
| • How does your organization and department deal with the changes you experience? | • What should the role of the organization / department be in this? |
| **Integration into practise**<br>• Looking back, what has changed since your first day in practice?<br>• What are the bottlenecks you are currently facing compared to your first day?<br>• Which uncertainties do you still face compared to your first day?<br>• Are you thinking about continuing working as a nurse even after the pandemic is over?<br>• What should change in healthcare to attract more former nurses? | • What is going better?<br>• Has a lot changed? How do you deal with this? What concrete steps should be taken to resolve these bottlenecks?<br>• Has a lot changed? How do you deal with this? What concrete steps should be taken to resolve these insecurities?<br>• Why yes/no?<br>• Why yes/no? |
| • What advice would you like to give to new returning nurses? |
| **Ending**<br>• Are there any important topics / points that we have not yet discussed and which you would like to address?<br>• Thanks for participating.<br>• Could I approach you again if new questions arise? |

## Appendix 3

Appendix three contains the coding scheme that was used to analyse data, categorised on themes, subcodes and codes.

Appendix 3 Appendix three contains the coding scheme that was used to analyse data, categorised on themes, subcodes and codes.
| **<u>Themes</u>** | **Description** | **Example** |
| --- | --- | --- |
| <u>Subcodes</u> |  |  |
| Codes |  |  |
| **<u>Theme: Job description and scope of practise</u>** |  |  |
| <u>Executing technical nursing activities.</u> | Segment in which returners discuss the agreements about taking up technical nursing activities. Including reserved actions. | *“There was someone who had wounds on her heels that had to be treated, but I was with someone who was from the nurse aide staff. He said, ‘I can't do that, someone from the level of nurse assistant staff should do that, he has to take care of those wounds.’ Yet that person said, ‘you are a nurse, if I tell you how to do it, can you do it?’ I thought* |
|  |  | *yes, this can't go wrong. It was nothing with injections or with drugs. I have been taking care of wounds for so long. So then I said, 'I will do it , but you should check it afterwards.' Well, I am competent, but actually I am not qualified. "* |
| <u>Clarity about role within the team</u> | Segments in which returners discuss whether their role was clear within their team or not. Including the expectations of the team members towards the returners. | *"Well, I got a call on a Tuesday asking if I wanted to come and work and if I could come within 48 hours. So I came there and their intention was that they immediately needed people who knew the ropes and could start working. While I came in with the idea, guys, I haven't done anything in healthcare for twenty years, help. I really want to do* |
|  |  | *something, but you have to say what the intention is and just give me instructions. “* |
| <u>Scope of practise</u> | Contains codes related to how the returners' role is ultimately executed regardless of their job description and how they experience this. | - |
| Comfortable / competent in role | Contains segments in which returners indicate that they felt competent in their role or to perform certain actions. | *“Well, I still feel like I have picked up things very quickly and could get started, I thought that was a very nice feeling. “* |
| Supporting role | Contains segments in which returners indicate that they perform a supporting role (regardless of the position in which they were previously hired). For example, some former nurses are hired as an | *“It was true that the nurses were ultimately responsible, but fortunately I never was alone on the ward after the first shift. So I was always with a nurse with a lot more* |
|  | independent nurse but ultimately perform in a more supportive role. Including reasons and experiences. | *experience. She was ultimately responsible, so I kept myself in the background a bit. "* |
| Defining boundaries | Segments in which returners indicate that they have to convey their own limitations regarding their competences | *"Uhm protect your boundaries, so know what belongs to your responsibility. And see if that is clear for the team, because otherwise you start with skewed expectations. "* |
| Logistical tasks | Segments in which returners indicate that (mainly in the beginning) they were also busy with logistics tasks due to the establishment of new departments. For example arranging the department, ordering missing materials, etc. | *"It was like this the first day for everyone, because what do we actually have? do we have a first aid kit, do we have an IV bag. You're trying to organize it. You try to supply the department and do all the logistics as well as possible.* |
| Grow in position | Segment in which returners indicate how they grow in | *"In the beginning it was a bit uncomfortable because I* |
|  | their role as their re-entry process progresses.<br>Including segments in which returners indicate that growth remains limited. | *have never done such physical work in a nursing home with people, and I did notice that I became a bit more comfortable with that towards the end. So that I just learned to really deliver physical care like that, so I learned from it. I am more skilled now than then. ”* |
| Proactive | Contains segments in which returners indicate that they should be proactive during the re-entry process. For example, when taking on tasks and asking questions to become more knowledgeable. | *“Everyone is busy with things that need to be arranged. Clinics have to open again, how should patient flow be organised, and you have a clinic that has to be moved outside, so they are now much more concerned with that. So if you end up in such a crisis situation and you are going to re-enter then you also need to have a proactive attitude and be capable to find your own way around* |
|  |  | *all this."* |
| Independent nurse | Segments in which returners report that they function as an independent nurse (regardless of the role in which returners were previously hired). Eg hired as a buddy, but eventually started working as an independent nurse.<br>Including reasons and experiences. | *"That they said within a day '(participants name), maybe you can just go back to work as an IC nurse. That might be a bit more convenient than as Buddy'. The next day I actually just started as an IC nurse. "* |
| Insecurities in role | Segments in which returners discuss insecurities within their role. | *"Yes I also felt a bit clumsy or something. I mean you have to adjust again. the last time I really washed someone's buttocks was in the nineties, so you know. So I thought... I felt a bit too .. are they really happy with that? Can I add value?"* |
| Nervousness about the unknown | Segments in which returners say that they find the new aspects of the department | *"Yes, it was very tense for everyone, it was the first week since the department* |
|  | and an unknown illness particularly tense. | *was established and everyone started working there, so it was new for everyone and everyone was a bit tense, because it was corona after all . “* |
| Tasks picked up quickly | Segments in which returners discuss the tasks they picked up quickly/easily. | *“Yes, that's especially when you're standing with the patient and you have to take care of the patient and you have to help the patient wash. You see how every nurse starts washing, and running, and communicating with the patients, and changing the beds. Those are actions you have done so many times and the observations you make, and the conversations you have at the bedside. Yes, you just pick these tasks up again and then you think, yes,* |
|  |  | *fortunately not much has changed. “* |
| Affinity with role | Segments in which returners express an affinity with caring and their role and are satisfied with their role. For example, working in healthcare again feels familiar. | *“As very positive and familiar. That sounds very strange, but I thought ‘oh dear, after 28 years suddenly I get back to such an organization and at the bedside. That is of course the reason why I once did my training. I really like that feeling; once a nurse always a nurse ”* |
| Time / space to learn within the role | Segments showing that returners needed a little more time to master certain tasks and/or realize that they cannot learn everything at once. | *I also noticed that when I walked through the corridors - well I have a fast pace anyway - but that the nurse said just walk a bit at a slower pace. Yes, you* |
|  |  | *have to give yourself time to get the hang of it again.* |
| Open attitude towards role | Segments showing that returners had an open and flexible attitude when they re-entered into care and had few expectations or requirements for their role.<br>"Just wanted to help" | *"Yes, and not anxious or anything. There just wasn't enough time to study everything very well. So, it was just like, 'go for it' and hope that I can patch it up if I misjudge it once. Yes, I can do that anyway, but you also have people who are very concerned about that. to do something wrong. I really thought, 'I will just do it and I know what I can do and I know what my limitations are'. That's how I work."* |
| **<u>Thema: Mental health</u>** |  |  |
| <u>Mental health impact</u> | Contains codes with segments about the impact of the re-entering process on the mental health of the returners. | - |
| Coping with death | Contains segments in which returners talk about the emotion of patients dying about corona or fear of dying of patients. | *“Yes, what I found very intense was that the manager also said, ‘yes gosh, Corona is a serious disease, because the situation of patients can deteriorate very suddenly and they can even die, which seems to be a very intense image’. So I was very afraid of that. Like oh dear, if that happens I might have that image of how someone literally suffocates in his own fluids on my mind for the rest of my life. ”* |
| Working with PPE | Segments in which returners say that working with PPE was demanding. | *“But it was continuously dressing and undressing, because for every room in and out you have to get your clothes on and off every time, so we were all the time getting dressed and* |
|  |  | *undressed. That was of course very labor-intensive and we used a lot of material. But it worked well, there was still enough material, so it was just a lot and very quickly responding to that, "* |
| Fear for infection | Segments in which returners talk about the fear of becoming infected themselves or the fear of infecting their family. | *"You know, when I got home I took a shower and only then I felt like I was really virus-free. I thought that was quite scary.* |
| Difficulties with consequences for patients' families | Segments in which returners indicate that they found it difficult that family was not allowed to visit the patient. | *"But you can get a little angry or frustrated yourself, that you think 'yes she will die soon and then their family just has not seen your mother again, is it really worth it. I found it very difficult, "* |
| Intensive | Segments in which returners indicate that their re-entering process was experienced as an intensive period. | *“Physical but also mentally, all the impressions... one day is easier than the other.. Yes, there are things that stick with you, like what you go through with people, families and the amount of information. You try to look things up yourself in the evening. So for me yes, but not too much, no not too much. It did not limit me. ”* |
| Letting go | Segments in which returners say it is important to let go of work. | *“I actually noticed that I can let go of this work more easily than when I'm still busy with all those managerial duties. I take that work home with me more often than the caregiving.* |
| Positive influence on mental health | Segments in which returners explain that the impact of their re-entering process on their mental health was actually positive or was not influenced at all. | *“Well not actually, well positive. Not negative anyway. It made me happy to be able to work again. Yes, you know I gained a valuable experience. I worked as a nurse again. but I did not impair my mental health.* |
| <u>Mental health support</u> | Codes with segments explaining the mental health support provided by the organizations and other support / coping mechanisms that re-entering nurses experienced / used. | - |
| Acknowledgement from environment | Segments in which returners speak about the acknowledgement and appreciation they have received from their environment and the | *“In (name city) you can nominate other people for a price, people who have done something good for the city. I got chocolates last week, which are* |
|  | organization in which they have worked. | *chocolates in the shape of a (characteristic of the city). I received that and we took a photo, which was posted on Facebook.* |
| Mental health support from manager | Segments in which returners report that team leaders / managers are vigilant about the mental health of employees | *“And then I thought maybe this is the time to say, ‘I have to take care of myself and I will go home to sleep’ and then a few days later I got a call from the team leader asking, ‘how are you and what happened’. Ehm, so they really pay attention to each other.”* |
| Media training | Segments about a media training that was set up by the business support team and its usefulness | *“Um well, I must say at first I was uncertain about how it would affect me. And because of the support that you get and the explanations on how things go, I only became less* |
|  |  | *insecure about it and I actually felt supported in the choices that I made. And that just helps me in daily life, because I know how to reduce the news and how to deal with everything that appears in the media and all the uncertainty. ”* |
| Psychological help | Segments in which returners say that there was a possibility to have a conversation with a psychologist or coach. | *“And I must also be honest, at (name of hospital) - and that is the case with many hospitals- it is all very well organised. Our psychologists said, ‘we are also available for our own employees’, and there was a consultation hour or people could call during the night. So people were really facilitated in this.”* |
| Talking with colleagues or | Segments in which returners | *"Also the contact with* |
| family | report that they sought support from colleagues and family. | *colleagues. You have a number of colleagues with whom you just notice that you can get along well, and um, you can talk to them."* |
| **<u>Theme: guidance and supervision</u>** | *Theme about how guidance and supervision is arranged and how returning nurses experience this.* | - |
| First point of contact / mentor | Segments from which it emerges whether returners are linked to a permanent mentor or / and that a first point of contact has been assigned. Including experiences and wishes. | *"Uhm and in practice I think it would be very good if someone was linked to a permanent mentor."* |
| Evaluation moment | Segments showing that returners make use of moments to evaluate or | *"Yes, that from time to time they have a conversation with that returner,* |
|  | whether they wish to do so. | *like 'gosh what problems do you actually encounter, what could we do differently and what could be improved?' ”* |
| Seeking guidance | Segments showing that returners themselves seek guidance and arrange supervision. | *“ In the beginning I was really linked to someone, when I started caring for patients independently, I always made sure that I had someone to go to, for myself, I arranged that myself. “* |
| Supporting role of manager | Segments in which returners speak about the manager's supporting role in the department. | *“Anyway, that's really ten points for our team management, because that's real. I do not know; no service is too crazy, you can request anything and they just see what they can do for you. “* |
| **<u>Thema: training</u>** |  |  |
| *<u>Received training</u>* | *Codes of segments related to* | - |
|  | *forms of training that returners have followed before or during their re-entry, including their experiences with this.* |  |
| Department-specific instructions | Contains segments in which returners talk about the instructions / introduction they have received about the department where they are placed. | *"What was expected of us, the rules, and someone came from the fire prevention department and from the GGD. So it was actually half a day. They showed us around, arranging uniforms, and everything."* |
| Practical training / Skills lab training | Contains segments in which returners talk about the practical training they have had. For example, a skills lab. | *"That was really very well organized together with the UT and the skills lab here in the hospital. Uhm, they started training people, so again the infusion pumps and learning things like that again. "* |
| COVID-19 specific training | Contains segments in which | *No, but we had very good* |
|  | returners talk about the instructions / training they have had focused on COVID-19. | *instructions about that.*<br>*Ehm, the red cross had arranged that for us. There were pamphlets hanging everywhere about the order in which you should take things off. And that nurse teacher, while we were busy with that briefing, she also gave good instruction. So I had that in mind, I knew that.”* |
| Self training | Contains segments in which returners report that they have sought out (extra) training themselves. | *“There is an e-learning that I just do and that helps enormously and that provides me support. I also check that COVID-19 site every day, because I think It's really great that they have also posted things about how to stay unharmed as a nurse. but where can you find protocols and e-learnings and things like* |
|  |  | *that.”* |
| E-learnings | Contains segments in which returners talk about the e-learnings they have followed. | *“There are several modules about for example resuscitation but also blood care. The organization has some kind of a learning environment where you are guided through the material and can do a practice test and eventually you can pass that thing”* |
| <u>No (full training) training</u> | Codes with segments showing that returners say that they have not received (full) training. | - |
| No training due to low complex care | Segments in which returners indicate that training was not necessary or not provided due to the low complexity of the tasks aligned with their position. | *“Uhm, I did not experience it as something I missed, uhm absolutely not, because it is just like you know, like cycling, knowledge on basic care is still there. I can do that ”* |
| No time for full training program | Segments in which returners indicate that there was no time to do a training before their return. | *"Actually, from the point of view of my nursing background, I think that there should be something, but we are now in a hectic situation in which I just see that a lot of people who work in practice are doing their utmost best. They actually also think that they should support more. "* |
| <u>Training needs</u> |  |  |
| Individual training programme | Segments in which returners discuss the importance of individualized training | *"Returners are people who often have quite some life-experience. It would be nice if training relates to this, so they do not have to do everything by default, but where you look at what someone needs. I have been out of the running for 28 years, but someone who was only absent for five years needs entirely something* |
|  |  | *else.”* |
| Practicing transfers/lifts | Segments in which returners indicate that they want to receive training on patient-lifts and transfers | *Uhm in this department - now again, I come back to that stupid patient lift (laughter) - I would have liked it if I would have known that from the beginning. I would have been able to work more independently and much faster”* |
| Practical training (need) | Segments in which returners express their desire for practical training. Including which practical actions they want to learn, eg reserved actions, pumps, etc. | *“For example, in terms of my reserved actions, I would have liked it if the things that had been changed or were adjusted in the protocol.. that someone would have demonstrated how to do those things right. Maybe I would have liked a skills lab, but it was not possible. ”* |
| COVID-19 related training | Segments in which returners | *“I would have wanted to* |
| (need) | indicate that they would have liked certain corona-related training. | *know especially a bit more about administering oxygen or also about the clinical picture itself. Um I might have wanted some more information about this."* |
| EPD training | Segments in which returners indicate that they would have liked to receive more training about the electronic patient file. | *"We should have invested a lot more time in that. We should have practiced with each other. We only went through some buttons on a powerpoint, like here you have the client, here you have the file, here you see this and here you see that. Well, of course that's not okay"* |
| Learning changed protocols /treatments | Segments showing that returners would have liked to receive some more guidance on changes in protocols and treatments. | *"Maybe discuss with someone which protocols are very common. For example, I had a patient with hyperglycemia and I* |
|  |  | *treated him in the way I thought I still had to, but the protocol had just been adjusted slightly. Anyway, if you are already on the work floor the next day, you do not have time to read 200 protocols. That just won't work. “* |
| **Theme: positive team dynamics** |  |  |
| <u>Positive team dynamic</u> |  |  |
| Team culture not yet established | Segments that show that there is no established team culture yet. Dynamics within the team have not yet been determined | *“So that makes it very different, everyone is a bit scanning, who are you, what can you do and what do you want. That is just completely different from entering an established group. I think if* |
|  |  | *we could have kept this up for a few more months, you would get that kind of dynamic, but it was not there yet."* |
| Team needs each other | Segments in which returners indicate that they had to work together because of the pressure of the crisis. | *"The team spirit, it was just almost fun. Just everyone was like 'maybe it will take a while and we just need you' and we needed them."* |
| Solidarity within the team | Segments showing that returners experience the team as willing to help. Solidarity towards each other. | *"Yes, everyone helps each other where they can. Everyone also puts the client first. I liked that so much, there were no hierarchical things or anything, everyone just did what they could, and everyone helped each other."* |
| Appreciation from the team | Segments in which returners indicate that they feel appreciation from their team | *"They were actually very happy with everything you could do and they really* |
|  |  | *emphasized that. I found that so surprising. Yes, I admitted that, it really felt very welcome ”* |
| Combatant team | Segments where returners indicate that the team is determined and combative to tackle the problems surrounding the pandemic together as a team. | *Ehm what I think is cool, that despite the whole new situation, despite all the hectic, I thought it was cool to discover that everyone had an enormous drive to get it all done, to get that job done. So everyone was very combative and all wanted to get involved.”* |
| Unity | Segments where returners indicate that they feel a great sense of unity within the team. Including wording such as: being part of the team, faces are in the same direction, close team, openness, etc. | *“And so a culture of cooperation arose, and suddenly there was no hierarchy anymore. People just started to consult with each other. There were things adapted in practice. People were open to any* |
|  |  | *comments. ”* |
| <u>Uninvolved in team</u> | Segments where returners describe moments when they felt less involved in the team. | *For example, a colleague said, ‘I am fine with taking care of that patient, but then I do want a good buddy.’ The room was filled with all the buddies, so when they picked someone, you knew that was a good buddy, and the rest was rubbish. You have to imagine that you are completely dressed up with a mask and eye protection, and then that is being said. I could not comprehend it.”* |
| <u>Varying team</u> | Codes with segments indicating that returners worked with a varying team and the consequences thereof | - |
| Estimating team | Segments showing that | *But it still took a while,* |
|  | returners and their colleagues had difficulty estimating what they could benefit from each other. For instance, in level of knowledge. | *because every time I was in a department with other colleagues, and everyone had a different background. So, we had to see who did what, but everyone was also very honest about that ‘I dare to do this, or I do not dare to do that, or I have not done that yet.’ Together we managed.* |
| New and unknown backgrounds | Segments showing that returners worked together with different colleagues from different backgrounds. | *“I also have to say that a number of those people are said secondment workers. So a department consists of only twenty permanent employees, such a small department the (name of hospital) has. Yes, the rest that was flown in from other departments or were secondment workers. “* |

## Appendix 4

In appendix four we elaborate on the BIG-law, reserved nursing procedures and legislation regarding nursing in the Netherlands during the COVID-19 pandemic.

To create clarity on the authorisation of medical procedures and to protect health professionals and patients from incompetence, the act on Professions in Individual Healthcare (‘BIG’ law) was formed (Wet Beroepen Individuele Gezondheid, 1993) in the Netherlands. This law includes a register of all qualified health professionals within individual healthcare and describes legal jurisdiction to execute specific medical activities (reserved procedures). Nurses acquire a BIG-registration when they have obtained a degree in nursing and must validate their registration every five years (ministerie van Volksgezondheid, Welzijn en Sport (VWS), 2020b; Wijmen et al., 1993). Validation can be done through the verification of at least 2080 hours of working experience within the individual healthcare in the previous five years or by passing a national re-registration exam. If nurses do not meet the requirements, they are no longer authorised to carry the title of a registered nurse, and all legal obligations and rights lapse (VWS, 2020b).

In the Netherlands, an existing grey area in the BIG law creates a possibility for former nurses to contribute to nursing during the COVID-19 pandemic. Registered nurses can delegate activities under specific conditions: nurses must guarantee proper supervision; be able to intervene if needed; and be able to assume that the person to whom the tasks are delegated, has the appropriate skills and knowledge (Wet Beroepen Individuele Gezondheid, 1993, §4, article 38).

Moreover, to accommodate the demand for extra qualified nurses during the pandemic within a short time-frame, the Dutch Ministry of Health made temporary adjustments in regulations (VWS, 2020a; Bruins, 2020). Firstly, all BIG re-register obligations for current health professionals were suspended until further notice. Secondly, former nurses whose BIG-registration expired after 1 January 2018 are temporarily allowed to work as a nurse without the requirements to re-register (VWS, 2020a; Bruins, 2020).

